# Estimated Glomerular Filtration Rate Slope and Kidney Outcomes in IgA Nephropathy

**DOI:** 10.1101/2025.06.15.25329659

**Authors:** Takaya Sasaki, Nobuo Tsuboi, Kentaro Koike, Hiroyuki Ueda, Masahiro Okabe, Shinya Yokote, Akihiro Shimizu, Keita Hirano, Tetsuya Kawamura, Takashi Yokoo, Yusuke Suzuki, the J-IGACS working group

## Abstract

**Background:** In IgA nephropathy (IgAN), early surrogate markers that reflect disease progression are critical for understanding long-term outcomes. While prior studies have suggested a potential role for eGFR slope, its validity as a surrogate endpoint requires further validation. We here investigated the association between longitudinal changes in eGFR and kidney outcomes in a large Japanese IgAN cohort.

**Methods:** Patients with biopsy-confirmed IgAN enrolled in the Japan IgA Nephropathy Cohort Study (J-IGACS) were analyzed. Individual eGFR slopes were estimated using a linear mixed-effects model. The association between eGFR slope and a composite kidney outcome, defined as a ≥40% decline in eGFR or the initiation of kidney replacement therapy, was assessed using joint modeling of longitudinal and survival data, adjusting for clinical and histopathological covariates, including current eGFR levels.

**Results:** During a median follow-up of 6 years in 937 patients (mean age 39 years; 51% male), 78 patients (8.3%) reached the composite kidney endpoint. Joint modeling analysis demonstrated that a steeper decline in eGFR slope was significantly associated with a higher risk of adverse kidney outcomes (hazard ratio per 1-sandard deviation slope decrease, 1.82; P<0.001). The association remained statistically significant in sensitivity analyses restricted to patients who had already received treatment at baseline, although the effect size was modestly attenuated.

**Conclusion:** Using joint modeling, we demonstrated that eGFR slope provides prognostic information beyond current eGFR levels in patients with IgAN. Our findings support the use of eGFR slope as a reliable surrogate endpoint for clinical trials and risk stratification in IgAN.

## Introduction

Although IgA nephropathy (IgAN) is recognized as the most common form of primary glomerulonephritis worldwide, predicting its long-term kidney outcomes remains a clinical challenge [1–3]. The disease is characterized by a typically slow, insidious, and heterogeneous progression [4–6], which complicates early risk stratification and often delays the initiation of timely, effective therapy. This persistent clinical uncertainty highlights an urgent need for early surrogate markers that can reflect disease activity and predict long-term kidney outcomes well before irreversible kidney damage occurs.

The estimated glomerular filtration rate (eGFR) slope offers significant theoretical advantages over cross-sectional assessments by providing a quantitative measure of kidney function decline over clinically meaningful timeframes [7, 8]. Unlike single time-point measurements, the eGFR slope captures the trajectory of kidney function deterioration, offering a more comprehensive understanding of disease progression. While numerous studies have investigated eGFR slope as a potential surrogate marker in IgAN, most of these have relied on meta-regression analyses of aggregated data [8–11], which provide important population-level insights but are inherently limited in evaluating individual-level longitudinal associations and within-patient variability over time, and direct validation in prospective well-characterized cohorts remains critically lacking. Such validation could strengthen the rationale for using eGFR slope as a surrogate endpoint for investigating this complex disease entity.

Recent advances in statistical methodology, particularly joint modeling of longitudinal biomarker trajectories and time-to-event outcomes, have enabled more rigorous examination of the association between biomarker trajectories (e.g., eGFR slope) and clinical events. Nonetheless, empirical evidence derived from large, well-characterized IgAN cohorts employing these advanced analytic methods remains scarce. Given the clinical and histopathological heterogeneity of IgAN, evaluating the utility and consistency of eGFR slope as a potential surrogate marker remains critically important.

To bridge this gap, we conducted a post hoc analysis of the Japan IgA Nephropathy Cohort Study (J-IGACS), a large, prospective registry of patients with biopsy-confirmed IgAN [12, 13]. Using joint modeling approaches, we examined the association between longitudinal changes in eGFR and clinically meaningful kidney outcomes, with the goal of evaluating eGFR slope as a candidate surrogate marker.

## Methods

### Study population

This study included patients with biopsy-proven IgAN who were enrolled in the Japan IgA Nephropathy Cohort Study (J-IGACS), a nationwide prospective cohort study [12, 13]. Participating facilities were encouraged to approach all eligible patients at each site, and a total of 1,130 patients provided written informed consent and were consecutively enrolled. Patients were eligible if they had a biopsy-confirmed diagnosis of IgAN, a kidney biopsy specimen containing at least 10 glomeruli, and at least two recorded eGFR measurements after enrollment. The population analyzed in the present study was derived from the original cohort by excluding individuals with missing data in any of the covariates used for multivariable adjustment in this analysis. Ethical approval was granted by The Jikei University School of Medicine (approval no. 37-069 [12706]). This study was conducted in accordance with the Declaration of Helsinki and reported in compliance with STROBE guidelines.

### Data collection

Clinical data were collected at the time of biopsy and subsequently at 6-month intervals, including age, sex, blood pressure, serum creatinine, estimated glomerular filtration rate, serum uric acid, 24-hour urinary protein excretion (the urinary protein-to-creatinine ratio was permitted in place of 24-hour urinary protein excretion at all time points except baseline), and urinary red blood cell count. eGFR was calculated using Uemura’s equation for individuals under 20 years of age [14] and Matsuo’s formula thereafter [15]. Kidney biopsy histopathological findings were scored using the Oxford MEST-C classification, based on standardized definitions for mesangial (M) and endocapillary (E) hypercellularity, segmental sclerosis (S), tubular atrophy/interstitial fibrosis (T), and crescents (C), in accordance with prior literature [16, 17].

### Exposure and outcome definition

The exposure was defined as annual eGFR slope from longitudinal eGFR measurements. Individual eGFR slopes, reflecting the rate of kidney function decline over time, were estimated using a linear mixed-effects model with random intercepts and random slopes. This approach allowed for the modeling of intra-individual variability and measurement timing differences across patients. The primary outcome was a composite kidney endpoint, defined as either a ≥40% decline from baseline eGFR or the initiation of kidney replacement therapy.

### Statistical analysis

Baseline patient characteristics were presented as mean (standard deviation [SD]) or median (interquartile range [IQR]) for continuous variables, and as frequencies and proportions for categorical variables.

Individual trajectories of eGFR were visualized using spaghetti plots. A generalized additive model was applied to assess potential non-linearity in the longitudinal eGFR trajectory. Based on these exploratory analyses, individual longitudinal changes in eGFR over time were characterized by using a linear mixed-effects model with random intercepts and random slopes, with eGFR as the dependent variable and follow-up time as the independent variable. This model incorporated all available repeated eGFR measurements for each participant and allowed estimation of subject-specific eGFR trajectories, including the underlying rate of eGFR change over time.

The association between the annual eGFR slope and the risk of the kidney composite endpoint was visualized using a scatter plot and a risk probability curve based on a logistic regression model. To further evaluate the relationship between longitudinal changes in eGFR and the risk of kidney outcomes, we employed joint modeling of longitudinal and survival data [18, 19]. This approach links the survival submodel to the subject-specific latent eGFR trajectory estimated from the linear mixed-effects model, rather than to a single observed eGFR slope, thereby integrating all repeated eGFR measurements and time-to-event data within a unified framework while accounting for individual variability.

Multivariable adjustments were made for the following baseline covariates: current eGFR value, age, log-transformed urinary protein excretion, the Oxford T score, use of RAAS inhibitors within one year after diagnosis, and steroid therapy within one year after diagnosis.

To assess the robustness of the primary findings, we conducted a sensitivity analysis restricted to patients who had received at least one disease-modifying treatment at baseline. Specifically, patients were included if they had a history of tonsillectomy, RAAS inhibitor use, or corticosteroid therapy prior to enrollment.

A two-sided P-value <0.05 was considered statistically significant. All statistical analyses were conducted using R version 4.5.0 (R Foundation for Statistical Computing, Vienna, Austria), except for the comparison of baseline characteristics between included and excluded patients, which was performed using SAS version 9.4 (SAS Institute Inc., Cary, NC, USA). Joint modeling was performed using the JMbayes2 package [19].

## Results

### Study population and eGFR slope estimation

After excluding 193 patients with missing data for any of the covariates from the initial 1,130 patients, a total of 937 patients with biopsy-confirmed IgAN (mean age 39; 51% male) were included in the present study. The baseline characteristics of the patients are presented in **Table 1**.

**Table 1.** Baseline characteristics in the present study.

|  | All patients<br>N = 937 |
| --- | --- |
| **Clinical characteristics** |  |
| Age, year, mean (SD) | 38.9 (16.7) |
| Male sex, n (%) | 481 (51.3) |
| Height, cm, mean (SD) | 161.8 (11.3) |
| Weight, kg, mean (SD) | 58.9 (13.7) |
| Body mass index, kg/m<sup>2</sup>, mean (SD) | 22.2 (3.8) |
| Systolic blood pressure, mmHg, mean (SD) | 122.1 (17.9) |
| Diastolic blood pressure, mmHg, mean (SD) | 73.9 (13.0) |
| eGFR, mL/min/1.73 m<sup>2</sup>, mean (SD) | 75.7 (28.8) |
| Uric acid, mg/L, mean (SD) | 58.4 (16.5) |
| UPE, g/day, median (IQR) | 0.575 (0.268–1.184) |
| Microhematuria*, n (%) | 799 (85.6) |
| **Pathological characteristics†** |  |
| M1, n (%) | 231 (29.2) |
| E1, n (%) | 332 (35.4) |
| S1, n (%) | 690 (73.6) |
| T1 or T2, n (%) | 202 (21.6) |
| C1 or C2, n (%) | 361 (38.5) |
| **Initial treatment** |  |
| RAAS inhibitor, n (%) | 533 (56.9) |
| Corticosteroid, n (%) | 603 (64.4) |
| Tonsillectomy, n (%) | 350 (44.2) |

Abbreviations: eGFR, estimated glomerular filtration rate; IQR, interquartile range; RAAS, renin-angiotensin-aldosterone system; SD, standard deviation; UPE, urinary protein excretion.
\*Defined as 5 or more red blood cells / high-power field.
†Indicates Oxford MEST-C scores: M, mesangial hypercellularity; E, endocapillary hypercellularity; S, segmental sclerosis; T, tubular atrophy/interstitial fibrosis; and C, cellular/fibrocellular crescents.

A comparison of baseline characteristics between included and excluded patients is provided in **Supplementary Table S1**. Overall, most clinical and pathological characteristics were comparable between the two groups. Although included patients were more likely to have received disease-directed therapy, other baseline characteristics were largely similar.

We analyzed a total of 10,543 longitudinal measurements of eGFR. The fitted curve based on a generalized additive model (**Figure 1A**) showed a generally steady decline in eGFR over time. After fitting a linear mixed-effects model with random intercepts and random slopes using eGFR as the dependent variable and follow-up time as the independent variable, individual eGFR slopes were obtained. The distribution of the estimated annual eGFR slopes was approximately normal, with a mean of −1.21 mL/min/1.73 m^2^/year (SD: 0.31), as shown in the histogram (**Figure 1B**).

**Figure 1.**
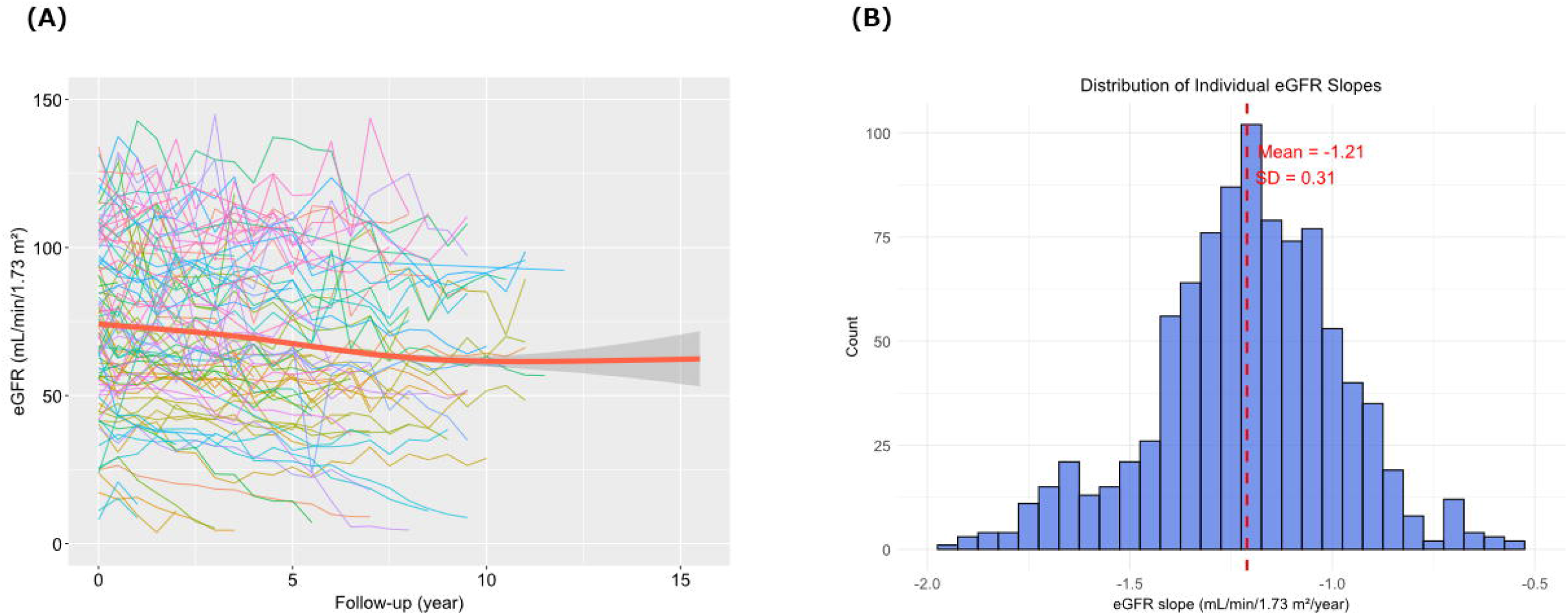
Longitudinal trends and distribution of estimated glomerular filtration rate (eGFR) slopes over time. (**A**) Time-course changes in eGFR during the entire observation period are plotted with a fitted curve calculated from a generalized additive model. For visualization purposes, individual trajectories (spaghetti plots) are shown for 100 randomly selected patients. (**B**) Histogram depicting the distribution of individual eGFR slopes calculated from the linear mixed-effects model.

### Association Between eGFR Slope and Kidney Outcomes

During the median follow-up of 6 years (IQR: 3–8), 68 patients (8.6%) reached the composite kidney endpoint, defined as either a ≥40% decline in eGFR from baseline or initiation of kidney replacement therapy. **Figure 2** shows the distribution of risk based on the eGFR slope and the fitted curve. eGFR slope was significantly negatively associated with the kidney composite endpoint (P <0.001). Next, a joint modeling analysis integrating longitudinal eGFR trajectories with time-to-event data was performed. Even after adjustment for baseline covariates—namely, current value of eGFR, age, log transformed urinary protein excretion, Oxford T score, use of RAAS inhibitors within one year and steroid therapy within one year—the joint modeling analysis revealed a significant association between steeper eGFR decline and increased risk of reaching the composite kidney endpoint (hazard ratio: 1.82 [95% confidence interval: 1.19–4.94] per 1-SD decrease in annual eGFR slope; P = 0.006; **Figure 3**). Results for other covariates are summarized in **Supplementary Table S2**.

**Figure 2.**
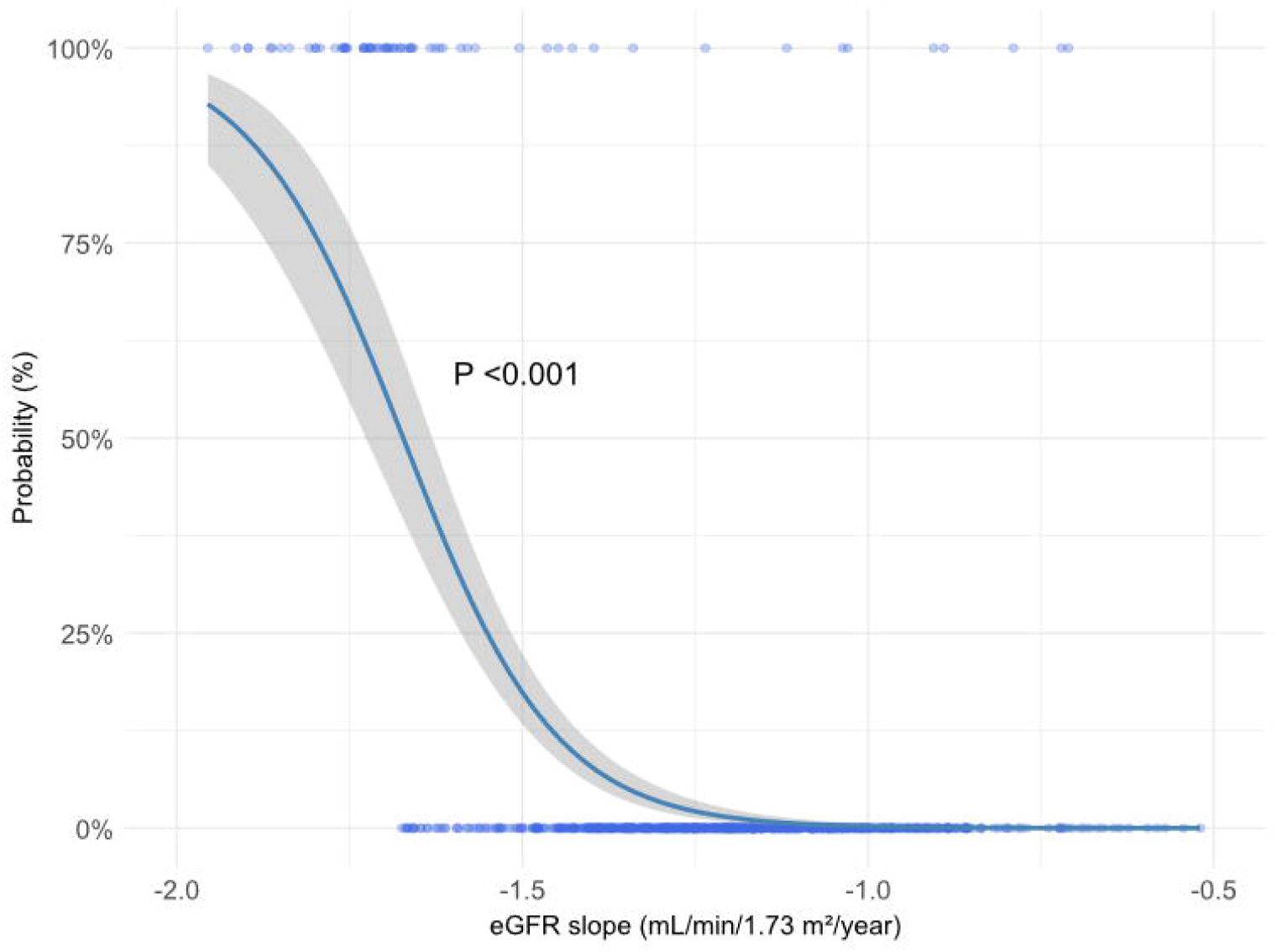
Relationship between the estimated glomerular filtration rate (eGFR) slope and risk of the kidney composite endpoint. Each dot represents an individual patient; the blue curve with grey shade represents the modeled event probability with 95% confidence interval estimated using logistic regression.

**Figure 3.**
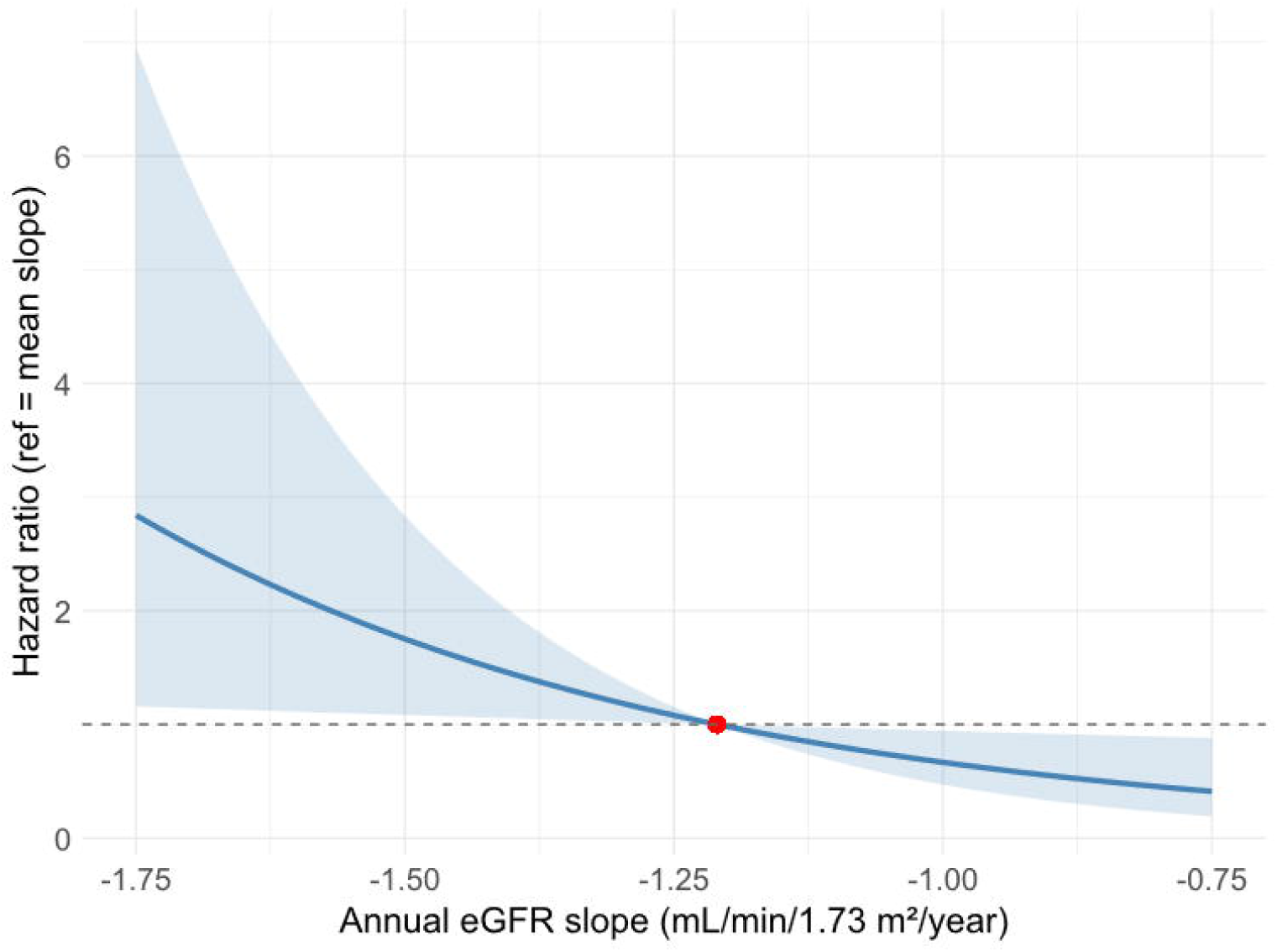
Association between the annual estimated glomerular filtration rate (eGFR) slope and kidney composite endpoint assessed using a joint modeling approach. Adjusted hazard ratios for the kidney composite endpoint are plotted against the annual eGFR slope estimated from joint modeling of longitudinal and survival data. The blue line represents the fitted hazard-ratio curve, and the shaded area indicates the 95% confidence interval. The red dot marks the reference point at an eGFR slope of −1.21 mL/min/1.73 m^2^/year, which is the mean value of eGFR slope.

### Sensitivity Analyses

In the sensitivity analysis restricted to patients who had received at least one treatment at baseline—defined as prior tonsillectomy, RAAS inhibitor use, or corticosteroid therapy—a steeper decline in eGFR remained significantly associated with a higher risk of the composite kidney endpoint, although the magnitude of the effect was attenuated compared with the primary analysis. Detailed results are presented in **Supplementary Table S3**.

## Discussion

In this large, well-characterized prospective cohort of patients with biopsy-confirmed IgAN, we applied joint modeling, integrating a linear mixed-effects model with survival analysis, to assess the association between longitudinal eGFR decline and time-to-event outcomes. This approach revealed a significant association between annual eGFR slope and adverse kidney outcomes, supporting its utility as a surrogate marker. Incorporating eGFR slope into clinical trial frameworks could meaningfully accelerate therapeutic development, enhance early intervention strategies, and ultimately contribute to improving long-term kidney outcomes in IgAN, a complex and heterogeneous disease.

Early and reliable surrogate markers are essential for evaluating treatment efficacy in IgAN, given its slow and heterogeneous progression [4–6]. Reliance on definitive kidney endpoints such as kidney failure necessitates prolonged follow-up and may delay recognition of treatment effects during earlier, potentially modifiable disease stages. While eGFR decline is intuitively linked to kidney prognosis, our use of joint modeling allowed us to disentangle the prognostic contribution of the rate of eGFR decline from that of the current eGFR value. Importantly, even after adjustment for the contemporaneous eGFR level, a steeper eGFR slope remained significantly associated with adverse kidney outcomes, indicating that longitudinal changes in kidney function convey prognostic information beyond cross-sectional measurements alone. This supports the concept that eGFR slope captures ongoing disease activity that may not be fully reflected by current absolute eGFR values, highlighting the clinical relevance of assessing prior eGFR slope.

Importantly, the significant association between a steeper decline in eGFR and an increased risk of reaching the composite kidney endpoint remained consistent in sensitivity analyses restricted to patients who had already received disease-modifying treatment at baseline. Although the magnitude of the association was modestly attenuated in this treated population, the direction and statistical significance of the relationship were preserved, underscoring the robustness of the primary findings. These results suggest that longitudinal eGFR decline retains prognostic value even among patients already receiving active therapy, likely reflecting residual disease activity beyond treatment effects. The consistency of this association supports the clinical relevance of eGFR slope as a stable and reproducible biomarker for disease progression and kidney outcomes in patients with IgA nephropathy.

Our results align with and extend prior research suggesting the utility of eGFR slope in chronic kidney disease populations [7–9]. While previous studies on IgAN have explored the association between eGFR slope and kidney outcomes using meta-regression analyses [9–11], this approach does not directly assess the individual-level association. In contrast, our use of joint modeling to simultaneously assess longitudinal changes in eGFR and survival outcomes allowed us to directly evaluate the association between eGFR slope and kidney outcomes within the complex pathology of IgAN. Traditional two-step approaches, modeling slope and survival separately, have been limited by time-lag bias and the assumption of a fixed eGFR slope, and thus have modeled the association with limited accuracy. By leveraging the entire longitudinal dataset, joint modeling provides a more accurate assessment of the association between eGFR changes and clinical outcomes.

Several limitations of this study should be acknowledged. First, although eGFR was measured at standardized six-month intervals, some degree of measurement variability and laboratory-related error is unavoidable and may have introduced noise into individual slope estimation. In addition, modeling eGFR decline as a linear process represents a simplifying assumption; while exploratory analyses using generalized additive models did not suggest pronounced non-linear patterns at the population level, alternative modeling strategies, such as spline-based or polynomial approaches, could influence the magnitude of effect estimates. Second, patients with missing covariate data were excluded from the analysis, which may have introduced selection bias. Although baseline clinical and pathological characteristics were largely comparable between included and excluded patients, included patients were more likely to have received prior treatments, and this imbalance should be considered when interpreting the results. Third, the generalizability of our findings to non-Japanese or more ethnically diverse populations remains to be established. In Japan, widespread urine screening facilitates earlier detection of IgAN [5], which may enhance the apparent utility of surrogate markers but limit applicability to settings with different disease phenotypes and treatment practices. Finally, as with any observational study, residual confounding cannot be completely excluded despite comprehensive multivariable adjustment.

In conclusion, our study provides robust evidence of a significant association between eGFR slope and adverse kidney outcomes in IgAN. By capturing longitudinal trajectories in kidney function, the eGFR slope was suggested to have utility as a marker of disease activity and practical surrogate endpoint. Its incorporation into clinical trial and clinical patient care may support earlier assessment of therapeutic intervention and inform disease monitoring strategies in patients with IgAN.

## Supplementary Material

**Supplementary Table S1. Comparison of baseline clinical and pathological characteristics between included and excluded patients**

**Supplementary Table S2. Joint modeling results for the association between eGFR slope and kidney outcomes in all patients**

**Supplementary Table S3. Joint modeling results for the association between eGFR slope and kidney outcomes in treated patients**

**Supplementary Item S1. Investigators list of the J-IGACS working group. Supplementary Item S2. STROBE Checklist**

Supplementary information is available at KI Report’s website.

## Authors’ Contributions

Conceptualization: Takaya Sasaki, Nobuo Tsuboi, Takashi Yokoo, Yusuke Suzuki.

Data curation: Takaya Sasaki.

Formal analysis: Takaya Sasaki.

Funding acquisition: Yusuke Suzuki.

Investigation: Takaya Sasaki, Nobuo Tsuboi.

Methodology: Takaya Sasaki.

Project administration: Nobuo Tsuboi, Takashi Yokoo, Yusuke Suzuki.

Supervision: Takashi Yokoo, Yusuke Suzuki.

Visualization: Takaya Sasaki.

Writing – original draft: Takaya Sasaki, Nobuo Tsuboi.

Writing – review & editing: Kentaro Koike, Hiroyuki Ueda, Masahiro Okabe, Shinya Yokote, Akihiro Shimizu, Keita Hirano, Tetsuya Kawamura, Takashi Yokoo, Yusuke Suzuki.

## Acknowledgments

This study was approved by the ethics review board of the Jikei University School of Medicine (approval no. 37-069 [12706]).

## Data availability statement

All data produced in the present study are available upon reasonable request to the authors.

