## Supplemental materials for "Estimated Glomerular Filtration Rate Slope and Kidney Outcomes in IgA Nephropathy"

### Supplementary Material

**Supplementary Table S1. Comparison of baseline clinical and pathological characteristics between included and excluded patients**
**Supplementary Table S2. Joint modeling results for the association between eGFR slope and kidney outcomes in all patients**
**Supplementary Table S3. Joint modeling results for the association between eGFR slope and kidney outcomes in treated patients**
**Supplementary Item S1. Investigators list of the J-IGACS working group**
**Supplementary Item S2. STROBE Checklist**

**Table S1. Comparison of baseline clinical and pathological characteristics between included and excluded patients**

| | No. of missing value of excluded patients | Excluded patients N=193 | Included patients N = 937 | P value |
|---|---|---|---|---|
| **Clinical characteristics** | | | | |
| Age, year, mean (SD) | 0 | 39.7 (16.2) | 38.9 (16.7) | 0.55 |
| Male sex, n (%) | 0 | 87 (45.1) | 481 (51.3) | 0.11 |
| Height, cm, mean (SD) | 32 | 163.8 (10.4) | 161.8 (11.3) | 0.031 |
| Weight, kg, mean (SD) | 47 | 59.3 (13.7) | 58.9 (13.7) | 0.023 |
| Body mass index, kg/m$^2$, mean (SD) | 51 | 22.7 (3.9) | 22.2 (3.8) | 0.55 |
| Systolic blood pressure, mmHg | 88 | 125.9 (17.3) | 122.1 (17.9) | 0.038 |
| Diastolic blood pressure, mmHg | 88 | 75.9 (13.0) | 73.9 (13.0) | 0.11 |
| eGFR, mL/min/1.73 m$^2$, mean (SD) | 28 | 75.2 (28.7) | 75.7 (28.8) | 0.85 |
| Uric acid, mg/L, mean (SD) | 39 | 60.4 (16.2) | 58.4 (16.5) | 0.17 |
| UPE, g/day, median (IQR) | 37 | 0.581 (0.296–1.110) | 0.575 (0.268–1.184) | >0.9 |
| Microhematuria*, n (%) | 0 | 135 (70.0) | 799 (85.6) | <0.001 |
| **Pathological characteristics†** | | | | |
| M1, n (%) | 79 | 29 (25.4) | 231 (29.2) | 0.45 |
| E1, n (%) | 79 | 33 (29.0) | 332 (35.4) | 0.17 |
| S1, n (%) | 79 | 89 (78.1) | 690 (73.6) | 0.31 |
| T1 or T2, n (%) | 79 | 25 (21.9) | 202 (21.6) | 0.93 |
| C1 or C2, n (%) | 79 | 39 (34.2) | 361 (38.5) | 0.37 |
| **Initial treatment** | | | | |
| RAAS inhibitor, n (%) | 0 | 90 (46.3) | 533 (56.9) | 0.009 |
| Corticosteroid, n (%) | 0 | 94 (48.7) | 603 (64.4) | <0.001 |
| Tonsillectomy, n (%) | 0 | 58 (30.1) | 350 (44.2) | <0.001 |

Abbreviations: eGFR, estimated glomerular filtration rate; IQR, interquartile range; RAAS, renin-angiotensin-aldosterone system; SD, standard deviation; UPE, urinary protein excretion.

*Defined as 5 or more red blood cells / high-power field.

†Indicates Oxford MEST-C scores: M, mesangial hypercellularity; E, endocapillary hypercellularity; S, segmental sclerosis; T, tubular atrophy/interstitial fibrosis; and C, cellular/fibrocellular crescents.

**Supplementary Table S2. Joint modeling results for the association between eGFR slope and kidney outcomes in all patients**

| Variable | Hazard ratio | 95% credible interval | P value |
|---|---|---|---|
| eGFR slope (per −0.31 mL/min/1.73 m²/year) | 1.82 | 1.17 – 3.14 | 0.0061 |
| Current eGFR (per 1 mL/min/1.73 m²) | 0.92 | 0.90 – 0.94 | <0.001 |
| Age (per 1 year) | 0.96 | 0.94 – 0.99 | 0.0007 |
| RAAS inhibitor use | 0.93 | 0.46 – 2.02 | 0.8334 |
| Corticosteroid use | 0.47 | 0.27 – 0.81 | 0.0105 |
| log-transformed UPE (per 1-log[g/day]) | 1.23 | 0.99 – 1.57 | 0.0628 |
| Oxford T score | 1.17 | 0.60 – 2.29 | 0.6475 |

Abbreviation: eGFR, estimated glomerular filtration rate; RAAS renin-angiotensin-aldosterone system; UPE, urinary protein excretion.

Footnote:

Hazard ratios were estimated using a joint model of longitudinal eGFR and time-to-event data.

The eGFR slope is expressed per 1-standard deviation decrease (−0.31 mL/min/1.73 m²/year).

**Supplementary Table S3. Joint modeling results for the association between eGFR slope and kidney outcomes in treated patients**

| Variable | Hazard ratio | 95% credible interval | P value |
|---|---|---|---|
| eGFR slope (per −0.31 mL/min/1.73 $m^2$/year) | 1.19 | 1.09 – 1.31 | 0.0002 |
| Current eGFR (per 1 mL/min/1.73 $m^2$) | 0.84 | 0.80 – 0.88 | <0.001 |
| Age (per 1 year) | 0.99 | 0.96 – 1.03 | 0.79 |
| RAAS inhibitor use | 1.16 | 0.06 – 64.5 | 0.99 |
| Corticosteroid use | 0.93 | 0.36 – 2.44 | 0.89 |
| log-transformed UPE (per 1-log[g/day]) | 1.27 | 0.90 – 1.83 | 0.18 |
| Oxford T lesion | 0.70 | 0.25 – 2.02 | 0.49 |

Abbreviation: eGFR, estimated glomerular filtration rate; RAAS renin-angiotensin-aldosterone system; UPE, urinary protein excretion.

Footnote:

Hazard ratios were estimated using a joint model of longitudinal eGFR and time-to-event data.

This sensitivity analysis was restricted to patients who had previously received at least one treatment at baseline, including tonsillectomy, RAAS inhibitor use, or corticosteroid therapy.

The eGFR slope is expressed per 1-standard deviation decrease (−0.31 mL/min/1.73 $m^2$/year).

**Supplementary Item S1. Investigators list of the J-IGACS working group**

**Chair:** Yusuke Suzuki (Department of Nephrology, Juntendo University Faculty of Medicine, Tokyo, Japan.)

**Co-chair:** Takashi Yokoo (Division of Nephrology and Hypertension, Department of Internal Medicine, The Jikei University School of Medicine, Tokyo, Japan.)

**Investigators:**

Ryosuke Aoki (Department of Nephrology, Juntendo University Faculty of Medicine, Tokyo, Japan.)

Shouichi Fujimoto (Department of Medical Environment Innovation, Faculty of Medicine, University of Miyazaki, Miyazaki, Japan.)

Yusuke Fukao (Department of Nephrology, Juntendo University Faculty of Medicine, Tokyo, Japan.)

Akihiro Fukuda (Department of Endocrinology, Metabolism, Rheumatology and Nephrology, Faculty of Medicine, Oita University, Oita, Japan.)

Akinori Hashiguchi (Department of Pathology, Keio University School of Medicine, Tokyo, Japan.)

Hiroshi Hataya (Department of Nephrology and Rheumatology, Tokyo Metropolitan Children's Medical Center, Fuchu, Tokyo, Japan.)

Keita Hirano (Division of Nephrology and Hypertension, Department of Internal Medicine, The Jikei University School of Medicine, Tokyo, Japan.)

Shiko Honma (Department of Pathology, The Jikei University School of Medicine, Tokyo, Japan.)

Daisuke Ichikawa (Division of Nephrology and Hypertension, Department of Internal Medicine, St. Marianna University School of Medicine, Kanagawa, Japan.)

Takafumi Ito (Department of Internal Medicine, Nephrology, Teikyo University School of Medicine, Teikyo University Chiba Medical Center, Chiba, Japan.)

Kensuke Joh (Department of Pathology, The Jikei University School of Medicine, Tokyo, Japan.)

Ritsuko Katafuchi (Kidney Unit, National Hospital Organization, Fukuoka-Higashi Medical Center, Fukuoka, Japan. Division of Nephrology, Medical Corporation Houshikai, Kano Hospital.)

Tetsuya Kawamura (Division of Nephrology and Hypertension, Department of Internal Medicine, The Jikei University School of Medicine, Tokyo, Japan.)

Masao Kihara (Department of Nephrology, Juntendo University Faculty of Medicine, Tokyo, Japan.)

Masao Kikuchi (Division of Cardiovascular Medicine and Nephrology, Department of Internal Medicine, Faculty of Medicine, University of Miyazaki, Miyazaki, Japan.)

Kentaro Koike (Division of Nephrology and Hypertension, Department of Internal Medicine, The Jikei University School of Medicine, Tokyo, Japan.)
Keiichi Matsuzaki (Department of Public Health, Kitasato University School of Medicine, Kanagawa, Japan.)
Kenichiro Miura (Department of Pediatric Nephrology, Tokyo Women's Medical University, Tokyo, Japan.)
Yoichi Miyazaki (Division of Nephrology and Hypertension, Department of Internal Medicine, The Jikei University School of Medicine, Tokyo, Japan.)
Takahito Moriyama (Department of Nephrology, Tokyo Medical University, Tokyo, Japan.)
Kumiko Muta (Advanced Medical Education Center, Nagasaki University School of Medicine, Nagasaki, Japan.)
Koichi Nakanishi (Department of Child Health and Welfare (Pediatrics), Graduate School of Medicine, University of the Ryukyus, Ginowan, Okinawa, Japan.)
Shinya Nakatani (Department of Metabolism, Endocrinology and Molecular Medicine, Osaka Metropolitan University Graduate School of Medicine, Osaka, Japan.)
Yoshihito Nihei (Department of Nephrology, Juntendo University Faculty of Medicine, Tokyo, Japan.)
Masako Nishikawa (Center for Research Promotion, The Jikei University School of Medicine, Tokyo, Japan.)
Tomoya Nishino (Department of Nephrology, Graduate School of Biomedical Sciences, Nagasaki University, Nagasaki, Japan.)
Ryoko Sakaguchi (Department of Pathology, The Jikei University School of Medicine, Tokyo, Japan.)
Takaya Sasaki (Division of Nephrology and Hypertension, Department of Internal Medicine, The Jikei University School of Medicine, Tokyo, Japan.)
Satoru Sanada (Department of Nephrology, Japan Community Healthcare Organization Sendai Hospital, Sendai, Japan.)
Sayuri Shirai (Division of Nephrology and Hypertension, Department of Internal Medicine, St. Marianna University School of Medicine, Kanagawa, Japan.)
Akihiro Shimizu (Division of Nephrology and Hypertension, Department of Internal Medicine, The Jikei University School of Medicine, Tokyo, Japan.)
Akira Shimizu (Department of Analytic Human Pathology, Nippon Medical School, Tokyo, Japan.)
Takanori Shibata (Division of Nephrology, Department of Medicine, Showa Medical University School of Medicine, Tokyo, Japan.)
Yuko Shima (Department of Pediatrics, Wakayama Medical University, Wakayama City, Wakayama, Japan.)
Hitoshi Suzuki (Department of Nephrology, Juntendo University Faculty of Medicine, Tokyo,

Japan.)
Kazuo Takahashi (Department of Biomedical Molecular Sciences, School of Medicine, Fujita Health University, Nagoya, Aichi, Japan.)
Nobuo Tsuboi (Division of Nephrology and Hypertension, Department of Internal Medicine, The Jikei University School of Medicine, Tokyo, Japan.)
Yasuhiko Tomino (Asian Pacific Renal Research Promotion Office, Medical Corporation SHOWAKAI, Shinjuku-ku, Tokyo, Japan.)
Hiroyuki Ueda (Division of Nephrology and Hypertension, Department of Internal Medicine, The Jikei University School of Medicine, Tokyo, Japan.)
Maki Urushihara (Department of Pediatrics, Institute of Biomedical Sciences, Tokushima University Graduate School, Tokushima, Tokushima, Japan.)
Takashi Yasuda (Naruse Kidney Clinic, Tokyo, Japan.)
Yoshinari Yasuda (Department of Advanced Science in Renal-Cardio Medicine/Nephrology, Gifu University Graduate School of Medicine, Gifu, Japan.)
Shinya Yokote (Department of Nephrology, Kawaguchi Municipal Medical Center, Saitama, Japan.)

**Supplementary Item S2. STROBE Checklist**

| | Item Description | Location (or reason for not reporting) |
|---|---|---|
| **Title and abstract** | | |
| 1a. Indicate the study's design | Indicate the study's design with a commonly used term in the title or the abstract. | Abstract Method |
| 1b. Abstract | Provide in the abstract an informative and balanced summary of what was done and what was found. | Abstract |
| **Introduction** | | |
| 2. Background / rationale | Explain the scientific background and rationale for the investigation being reported. | Introduction 1st-3rd paragraphs |
| 3. Objectives | State specific objectives, including any prespecified hypotheses. | Introduction 3rd paragraph |
| **Methods** | | |
| 4. Study design | Present key elements of study design early in the paper. | Method Study design and population |
| 5. Setting | Describe the setting, locations, and relevant dates, including periods of recruitment, exposure, follow-up, and data collection. | Method data acquisition, Exposure Definition and Propensity Score-based Weighting Approach |
| 6a. Eligibility criteria | **Cohort study:** Give the eligibility criteria, and the sources and methods of selection of participants. Describe methods of follow-up. **Case-control study:** Give the eligibility criteria, and the sources and methods of case ascertainment and control selection. Give the rationale for the choice of cases and controls. **Cross-sectional study:** Give the eligibility criteria, and the sources and methods of selection of participants. | Method Study design population |
| 6b. Matching criteria | **Cohort study:** For matched studies, give matching criteria and number of exposed | Not applicable |

| | | |
|---|---|---|
| | and unexposed. **Case-control study:** For matched studies, give matching criteria and the number of controls per case. | |
| 7. Variables | Clearly define all outcomes, exposures, predictors, potential confounders, and effect modifiers. Give diagnostic criteria, if applicable. | Method data acquisition, Exposure Definition and Propensity Score-based Weighting Approach |
| 8. Data sources / measurement | For each variable of interest give sources of data and details of methods of assessment (measurement). Describe comparability of assessment methods if there is more than one group. | Method data acquisition, Exposure Definition and Propensity Score-based Weighting Approach, |
| 9. Bias | Describe any efforts to address potential sources of bias. | Method Exposure Definition and Propensity Score-based Weighting Approach, statistical analysis |
| 10. Study size | Explain how the study size was arrived at. | Method Study population |
| 11. Quantitative variables | Explain how quantitative variables were handled in the analyses. If applicable, describe which groupings were chosen, and why. | Method data collection, Exposure Definition and Propensity Score-based Weighting Approach, Endpoints |
| 12a. Statistical methods | Describe all statistical methods, including those used to control for confounding. | Method statistical analysis |
| 12b. Statistical methods – subgroups and interactions | Describe any methods used to examine subgroups and interactions. | Method statistical analysis |
| 12c. Statistical methods – missing data | Explain how missing data were addressed. | Not applicable (complete data analysis) |
| 12di. Statistical methods – loss to follow-up | **Cohort study:** If applicable, describe how loss to follow-up was addressed. | Not applicable (post-hoc analysis) |

| | | |
|---|---|---|
| 12dii. Statistical methods – matching cases and controls | **Case-control study:** If applicable, explain how matching of cases and controls was addressed. | Not applicable |
| 12diii. Statistical methods – sampling strategy | **Cross-sectional study:** If applicable, describe analytical methods taking account of sampling strategy. | Not applicable |
| 12e. Statistical methods – sensitivity analyses | Describe any sensitivity analyses. | Method statistical analysis |
| **Results** | | |
| 13a. Participant numbers | Report the numbers of individuals at each stage of the study—e.g., numbers potentially eligible, examined for eligibility, confirmed eligible, included in the study, completing follow-up, and analysed; Consider use of a flow diagram. | Results<br>Baseline Characteristics |
| 13b. Participants – non-participation | Give reasons for non-participation at each stage. | Results<br>Baseline Characteristics |
| 13c. Participants – flow diagram | Consider use of a flow diagram. | We stated the number instead of using flow diagram |
| 14a. Descriptive data – participant characteristics | Give characteristics of study participants (e.g., demographic, clinical, social) and information on exposures and potential confounders. Present the information in a table. | Results<br>Baseline Characteristics |
| 14b. Descriptive data – missing data | Indicate the number of participants with missing data for each variable of interest. | Complete case analysis |

| | | |
|---|---|---|
| 14c. Descriptive data – follow-up time | **Cohort study:** Summarise follow-up time—e.g., average and total amount. | Kidney Outcomes, eGFR slope, proteinuria ratio |
| 15. Outcome data | **Cohort study:** Report numbers of outcome events or summary measures over time. **Case-control study:** Report numbers in each exposure category, or summary measures of exposure. **Cross-sectional study:** Report numbers of outcome events or summary measures. | Kidney Outcomes, eGFR slope, proteinuria ratio |
| 16a. Main results | Give unadjusted estimates and, if applicable, confounder-adjusted estimates and their precision (e.g., 95% confidence intervals). Make clear which confounders were adjusted for and why they were included. | Kidney Outcomes, eGFR slope, proteinuria ratio, Comparison with Meta-Regression Estimations |
| 16b. Main results – category boundaries | Report category boundaries when continuous variables were categorised. | Not applicable |
| 16c. Main results – risk | If relevant, consider translating estimates of relative risk into absolute risk for a meaningful time period. | Not applicable |
| 17. Other analyses | Report other analyses done—e.g., analyses of subgroups and interactions, and sensitivity analyses. | Comparison with Meta-Regression Estimations |
| **Discussion** | | |
| 18. Key results | Summarise key results with reference to study objectives. | Discussion 1st paragraph |
| 19. Limitations | Discuss limitations of the study, taking into account sources of potential bias or imprecision. Discuss both direction and magnitude of any potential bias. | Discussion 4th paragraph |
| 20. Interpretation | Give a cautious overall interpretation considering objectives, limitations, | Discussion 1st-4th paragraph |

| | | |
|---|---|---|
| | multiplicity of analyses, results from similar studies, and other relevant evidence. | |
| 21. Generalisability | Discuss the generalisability (external validity) of the study results. | Discussion 4th paragraph |
| **Other information** | | |
| 22. Funding | Give the source of funding and the role of the funders for the present study and, if applicable, for the original study on which the present article is based. | Funding |
